# Influence of Early Endomyocardial Biopsy on the Treatment and Prognosis in Fulminant Myocarditis

**DOI:** 10.64898/2026.09.24.26363980

**Authors:** Yasuyoshi Takei, Taishiro Chikamori, Koshiro Kanaoka, Kenji Onoue, Kazuhiro Satomi, Yoshihiko Saito

## Abstract

**BACKGROUND:** Because of the high mortality rate associated with fulminant myocarditis (FM), there is limited evidence regarding the prognostic efficacy of endomyocardial biopsy (EMB). In this study, we aimed to evaluate the influence of EMB in the early phase after admission on treatment and prognosis in FM using a large-scale nationwide registry in Japan.

**METHODS:** Patients with FM who required catecholamine or mechanical circulatory support between 2012 and 2017 from 235 hospitals across Japan were included. The primary outcome of death or heart transplantation within 90 days according to performance of early EMB were determined using Kaplan-Meier and Cox regression analyses. Factors associated with performance of early EMB, and subsequent medical treatment were analyzed.

**RESULTS:** The median age of 736 patients with clinical FM was 56 years, and 407 patients (55%) received EMB. Most EMB (89%) were performed during the early phase (within 5 days of admission) and 63% were performed on the first day of admission. The prognosis of FM patients who underwent EMB was more favorable than that of those who did not based on the landmark analysis (log-rank test, *P*=0.04). Factor analysis demonstrated that FM patients who underwent EMB had a lower left ventricular ejection fraction, higher incidence of cardiogenic shock on admission, and higher rate of early mechanical circulatory support, as well as early initiation of corticosteroids and guideline-directed medical therapy (GDMT) for heart failure. Absence of any GDMT was associated with a less favorable prognosis on multivariate analysis (hazard ratio, 4.34 [95% CI, 3.14–6.01]; *P*<0.0001).

**CONCLUSIONS:** Although performing EMB in the early phase after admission depends on disease severity in FM patients, it is associated with early initiation of advanced medical treatment and may contribute to improving the prognosis.

**WHAT IS NEW?:**

- Fulminant myocarditis patients who did not undergo endomyocardial biopsy had a higher mortality rate than those who underwent endomyocardial biopsy after considering immoral time bias associated with the performance of early endomyocardial biopsy.
- Early endomyocardial biopsy was associated with early initiation of mechanical circulatory support, early administration of corticosteroids and immunoglobulins and a higher rate of administration of guideline-directed medical therapy for heart failure during hospitalization.
- In the prognostic factor analysis, guideline-directed medical therapy for heart failure during hospitalization was strongly associated with a favorable prognosis of patients with fulminant myocarditis.

**WHAT ARE THE CLINICAL IMPLICATIONS?:**

- There is limited evidence regarding the influence of endomyocardial biopsy in the early phase after admission on the treatment and prognosis in patients with fulminant myocarditis. Our results provide valuable insights indicating that combination of accurate histological diagnosis by endomyocardial biopsy in the early phase and appropriate cardioprotective therapy leads to a favorable prognosis of patients with fulminant myocarditis.

---

Myocarditis is a myocardial inflammatory disease caused by viral infections, autoimmune diseases, and adverse drug reactions.^1^ Its clinical presentations range from asymptomatic cases with electrocardiographic changes to fatal cases attributable to severe heart failure or cardiogenic shock. Fulminant myocarditis (FM) is a rare phenotype of myocarditis characterized by hemodynamic collapse requiring inotropes or temporary mechanical circulatory support devices (MCS).^2,3^ Although current knowledge of FM in clinical settings remains limited, we previously reported a very high mortality rate of FM (approximately 30% at 90 days), and identified prognostic factors on the basis of pathohistological findings.^4^ Indeed, histological diagnosis by endomyocardial biopsy (EMB) is strongly recommended for FM because it is the only method that enables the accurate diagnosis and pathogenic characterization of myocarditis. Several previous multicenter studies have shown that histological diagnosis by EMB improves the prognosis of FM.^5,6^ However, how histological diagnosis by EMB, particularly in the early phase after admission, affects the medical treatment, management and prognosis of patients with FM has not been clarified to date. Therefore, in this study we investigated the influence of EMB in the early phase after admission on medical treatment and prognosis in patients with FM using the largest cohort study database available in Japan.

## METHODS

### Study Design and Population

The Japanese Registry of Fulminant Myocarditis is a nationwide multicenter, retrospective cohort study involving 235 cardiovascular training hospitals in Japan (Figure 1). The methods of data collection were reported previously.^4^ Using the Japanese Registry of All Cardiac and Vascular Diseases–Diagnosis Procedure Combination (JROAD–DPC) discharge database, which is a claims database covering more than 60% of all cardiovascular training hospitals in Japan,^7,8^ patients who were diagnosed as having myocarditis between April 2012 and March 2017 using the International Classification of Diseases–10 (ICD–10) codes I–40, I–41, or I–423 were extracted. These patients were categorized into FM and non-FM based on catecholamine or MCS use during their hospitalization.^9^ Individual patient data were collected from each facility if they agreed to join the registry and local institutional review board approval was obtained. Cardiologists at each participating hospital manually reviewed the charts to extract individual data. Histologically confirmed myocarditis was defined on the basis of the clinical diagnostic criteria of the European Society of Cardiology or the Japanese Circulation Society, and the histologic definitions of World Health

**Figure 1.**
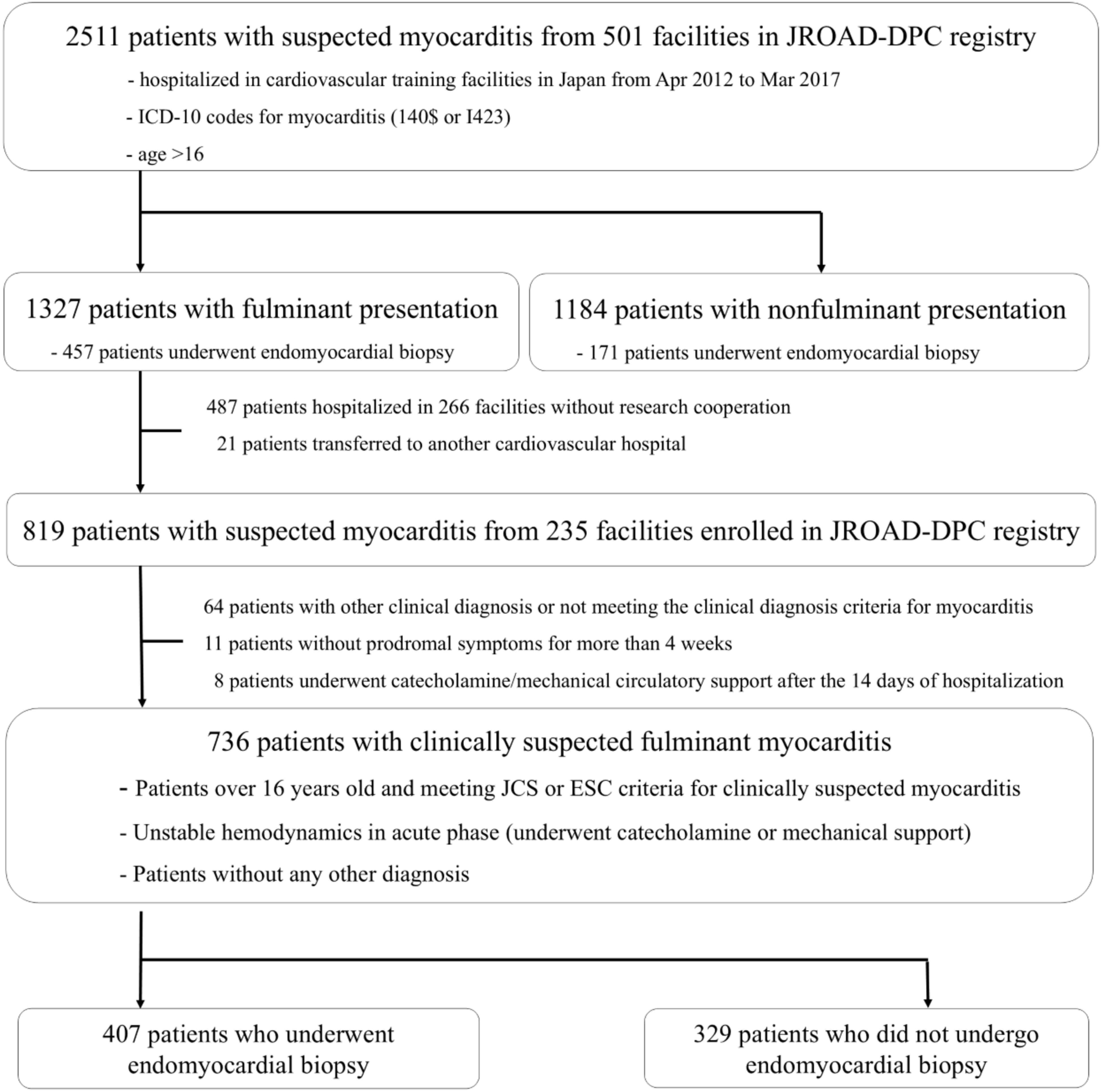
Patient enrollment and assignment flow chart. EMB, endomyocardial biopsy; ESC, European Society of Cardiology; ICD–10, International Classification of Diseases–10; ISFC, International Society and Federation of Cardiology; JCS, Japanese Circulation Society; JROAD–DPC, Japanese Registry of All Cardiac and Vascular Diseases–Diagnosis Procedure Combination; WHO, World Health Organization

Organization/International Society and Federation of Cardiology criteria.^10–12^ FM was defined on the basis of previous studies.^4,9^ Exclusion criteria included patients with previously known cardiac diseases, Takotsubo cardiomyopathy, sepsis-induced cardiomyopathy, peripartum cardiomyopathy, heart transplantation, and patients younger than 16 years. Patients were also excluded if they were diagnosed as having other diseases, if their symptoms had appeared more than 30 days before hospitalization, or they started to use catecholamine or MCS after the 14th day of hospitalization.

As baseline data, patients’ demographic factors, histories of myocarditis, autoimmune disease and atherosclerotic risk factors, type and timing of initial symptoms, electrocardiogram, echocardiographic parameters, and laboratory examination data at admission were collected. Data regarding major myocarditis-associated events, such as ventricular arrhythmia, high-degree advanced atrioventricular conduction abnormalities, and cardiogenic shock, as well as indications for MCS including intra-aortic balloon pumping (IABP), extracorporeal membrane oxygenation (ECMO), and ventricular assist device (VAD) use were also collected. In addition, information regarding the use of immunomodulatory therapies including intravenous corticosteroids, intravenous immunoglobulins and guideline-directed medical therapy (GDMT) for heart failure during hospitalization were collected.

To assess histological diagnosis using EMB specimens, experienced cardiovascular pathologists who were blinded to patient outcomes independently evaluated the biopsy specimens. To determine the prognostic efficacy of histological diagnosis by EMB and its influence on subsequent medical treatment, FM patients were divided into those with and those without histological diagnosis by EMB. The primary outcome was all-cause mortality or heart transplantation (HTx) within 90 days of hospital admission. Follow-up data were obtained through medical records or telephone interviews. This study was conducted in accordance with the principles of the Declaration of Helsinki. The study protocol was approved by the Ethics Committees of Nara Medical University (study registration number: 2256) in July 2019, the Japanese Circulation Society (study registration number: 10) in November 2019 and Tokyo Medical University (study registration number: T2020-0041). This study was registered at the University Hospital Medical Information Network Clinical Trials Registry of Japan (registration number: UMIN000039763).

### Statistical Analysis

Continuous variables were expressed as the median and 25th to 75th interquartile range (IQR) and compared using the unpaired Student *t*-tests, and categorical variables were expressed as percentages for baseline clinical characteristics. The Wilcoxon rank-sum and Pearson chi-squared tests were used to compare continuous and categorical variables between FM patients with histological diagnosis by EMB and those without. The Kaplan-Meier method with the log-rank test and Cox regression models were used to analyze the incidence of death or HTx within 90 days of hospital admission. All statistical tests were 2-sided and *P* values less than 0.05 were considered to indicate a statistically significant difference between groups. SPSS version 29.0 software (SPSS, Inc., Chicago, IL, USA) was used for all statistical analyses.

## RESULTS

### Study Population and Characteristics

A total of the 2511 patients with suspected acute myocarditis in the JROAD–DPC database (1327 with fulminant presentation and 1184 without fulminant presentation), who were hospitalized between April 2012 and March 2017 were extracted. Of the 1327 patients extracted as having FM, 819 patients hospitalized in 235 cardiovascular facilities with permission to join this project were included in this study. Of the 736 patients with clinical features of FM, 329 patients did not undergo EMB, and 407 patients who underwent EMB were finally included in the study (Figure 1). Furthermore, of all the patients who underwent EMB, 63.3% underwent EMB on the first day of admission and 89.4% underwent EMB in the early phase, i.e., within 5 days of admission (Figure 2). Demographic data of the 736 patients are shown in Table 1. The median (IQR) age was 56 (39–67) years, the most frequent predominant prodromal symptom was fever (61.5%) and 77.1% patients had cardiogenic shock on hospital admission. The median (IQR) value of left ventricular ejection fraction and brain natriuretic peptide were 30 (20–44) % and 577 (294–1120) pg/mL, respectively. Most patients with FM (97.4%) were admitted to the facilities with cardiovascular specialists certified by the Japanese Circulation Society, with 65.2% of patients receiving MCS within 2 days of admission, and 78.3% of patients receiving them within 5 days of admission.

**Figure 2.**
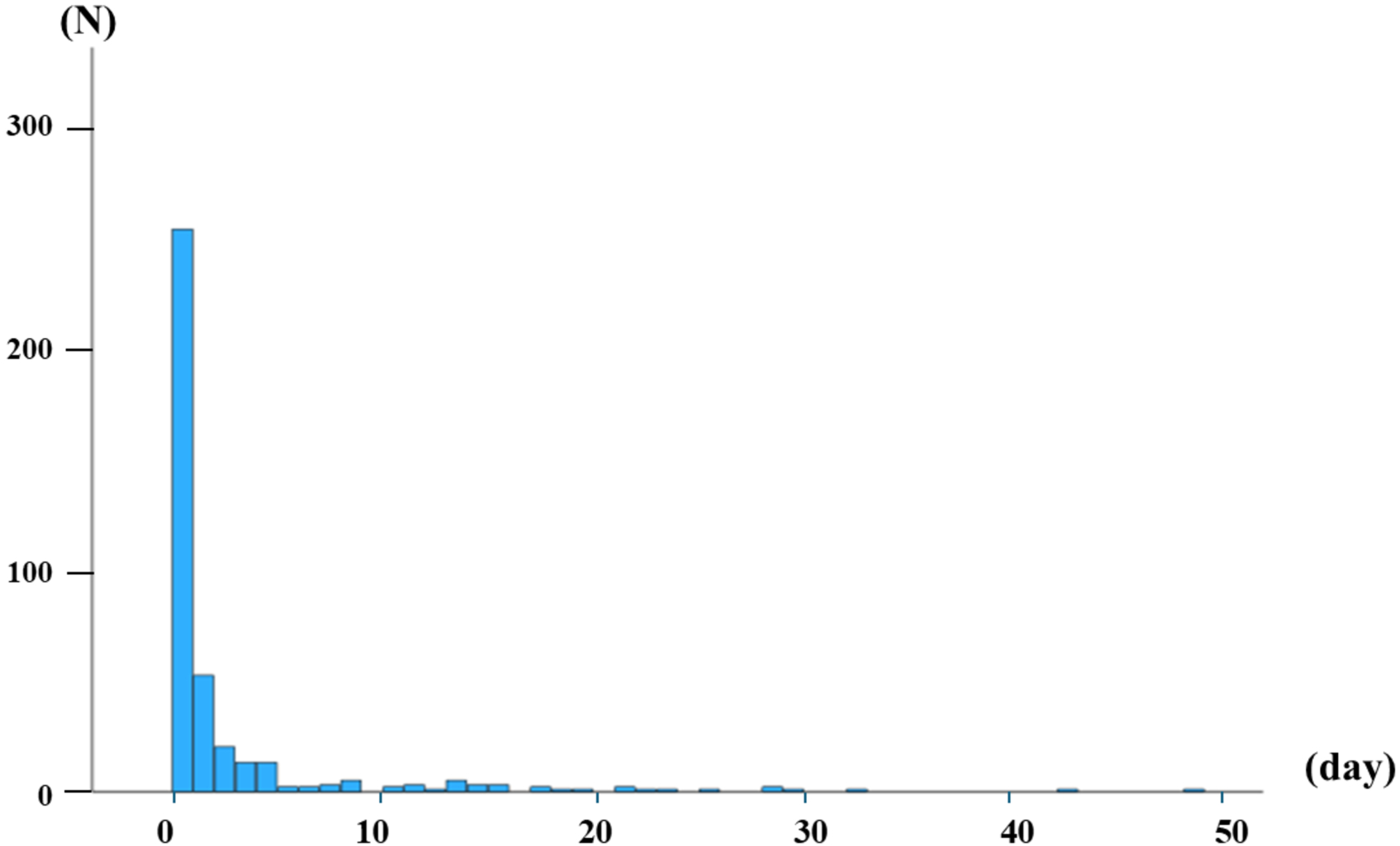
Distribution of dates of performance of endomyocardial biopsy. The rates of myocardial biopsy performance were 63.3% on the first day, 76.2% within two days, and 89.4% within five days of admission.

**Table 1.**
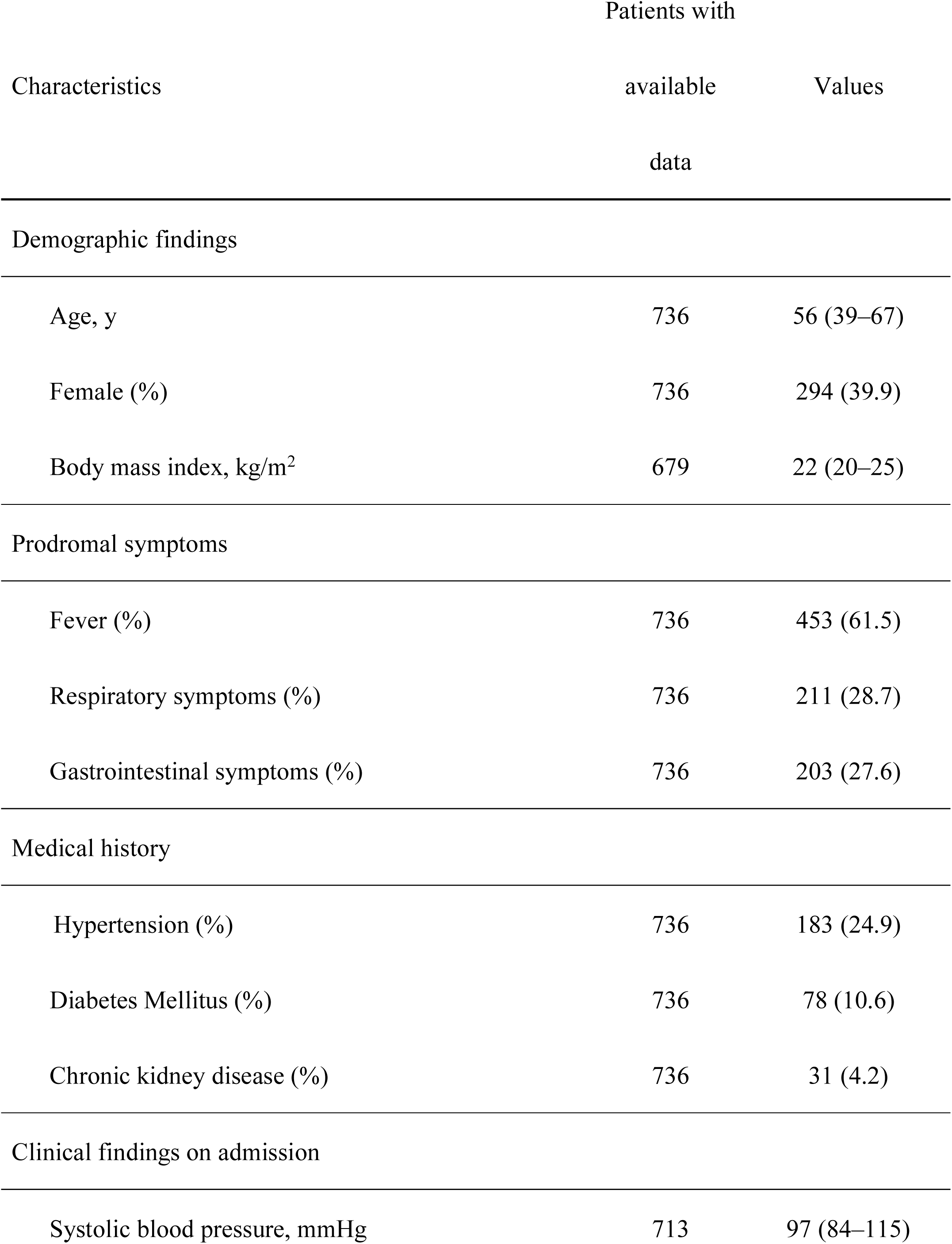
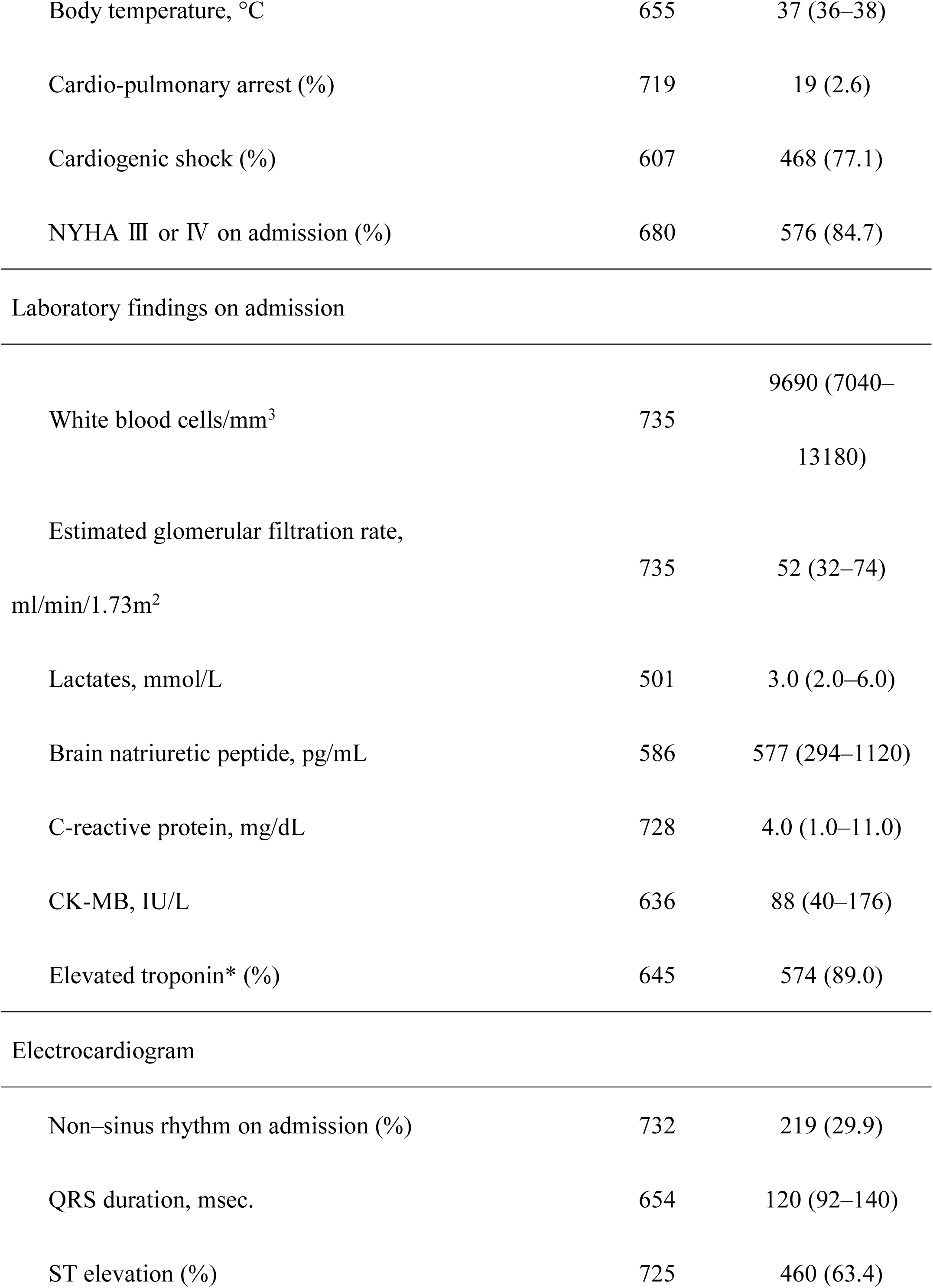
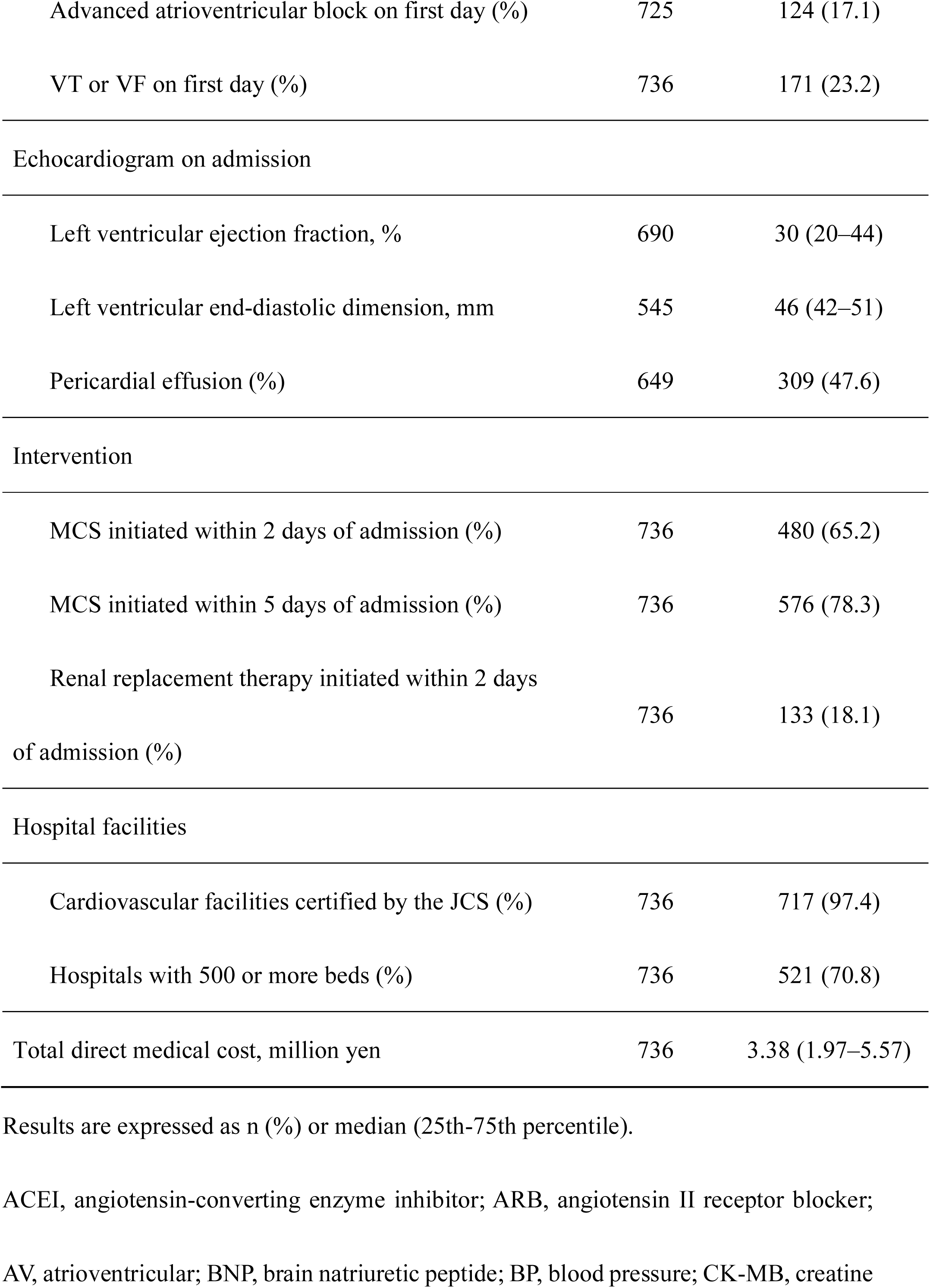
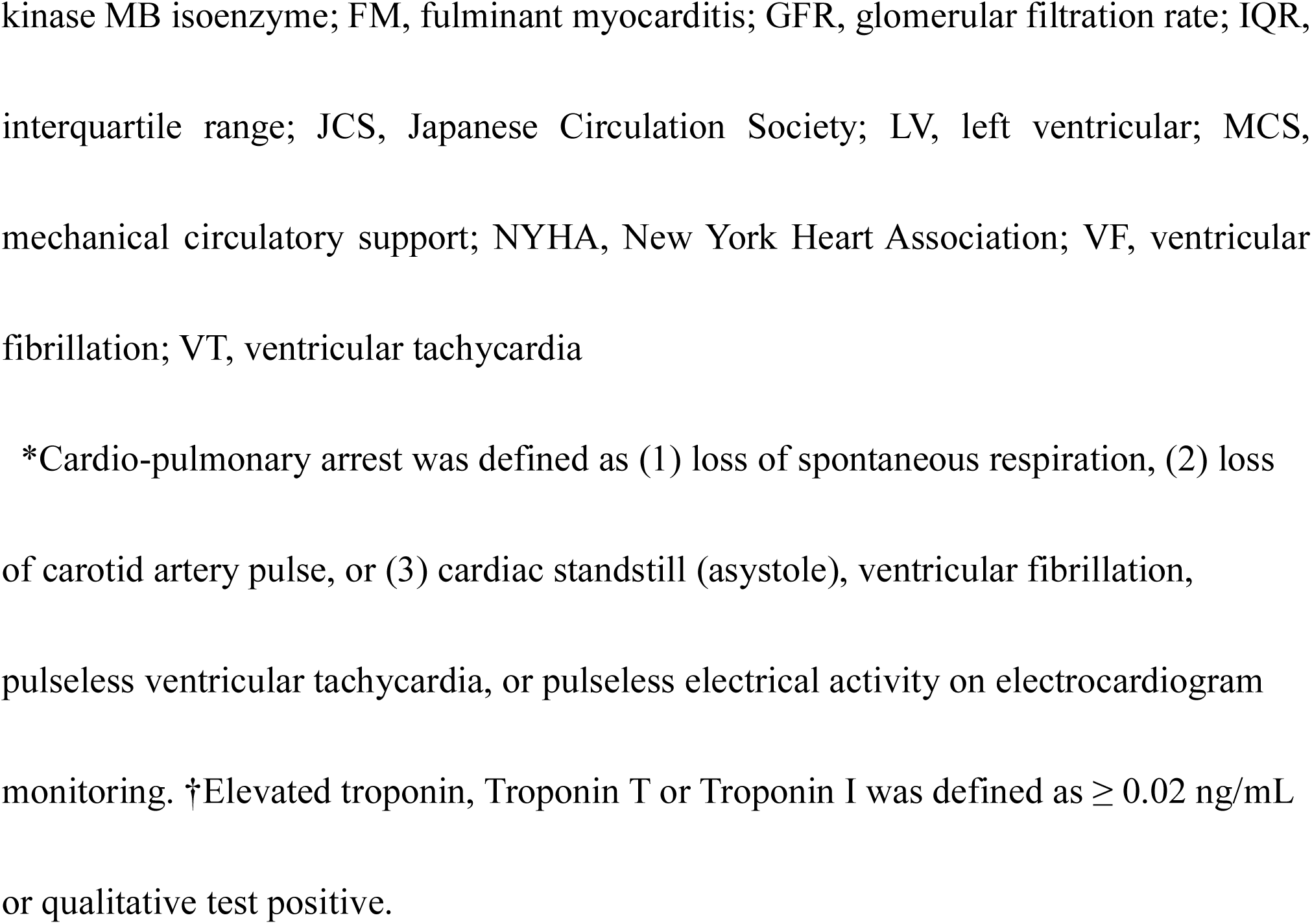
Baseline Characteristics in Patients with FM.

### Ninety-day Mortality Analysis

To compare the 90-day mortality between FM patients with EMB and those without, we first performed a crude analysis using the Kaplan-Meier and the log-rank methods (Figure 3). The prognosis was more favorable in FM patients with EMB than those without. However, because the Kaplan-Meier curves confirmed a difference between the 2 groups in the very early phase from admission, we calculated the proportion of deaths within 5 days of admission, as approximately 90% of EMB procedures were completed within this period. Of all FM patients who died, 81 (28.5%) died within 5 days of admission. The mortality rate for the FM patients with EMB group within 5 days of admission was 6.1% (25/407 patients), whereas that for the FM without EMB group was 17.0% (56/329 patients). The chi–squared test showed a *P* value of <0.0001, indicating a significantly higher mortality rate in the latter group. This result indicates the presence of immortal time bias in the crude comparison of mortality rates between the 2 groups. Therefore, we performed a landmark analysis in patients with FM who were confirmed to be alive on the fifth day of admission, categorizing those who underwent EMB within 5 days of admission to the FM patients with EMB group and those who did not to the FM patients without EMB group. After excluding the 81 patients who died within 5 days of admission, among the remaining 655 patients with FM, 90-day mortality was more favorable in FM patients with EMB than in those without EMB using the landmark analysis (Figure 4).

**Figure 3.**
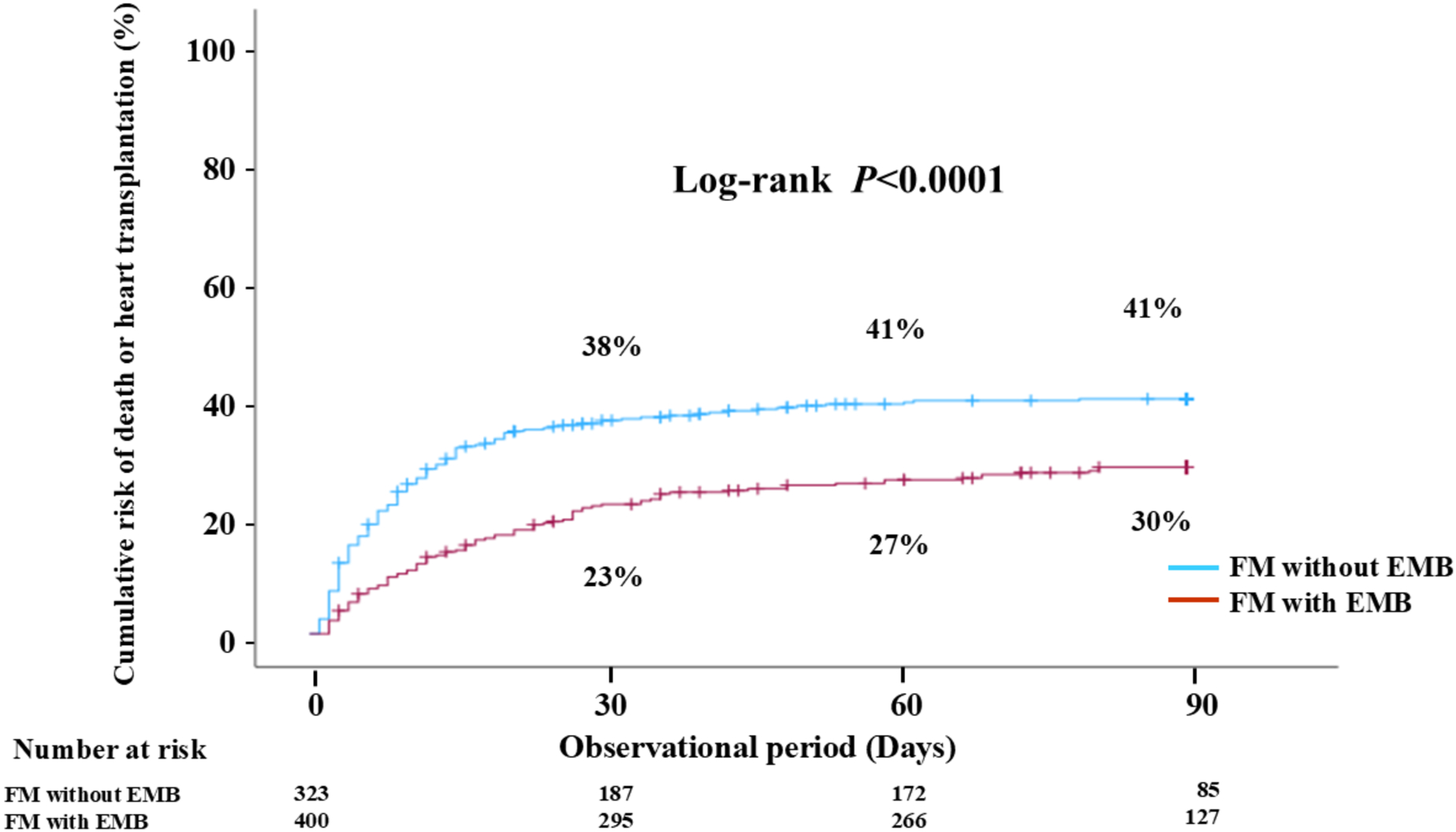
Crude 90-day prognostic analysis for death or heart transplantation according to endomyocardial biopsy in patients with fulminant myocarditis. FM, fulminant myocarditis; EMB, endomyocardial biopsy.

**Figure 4.**
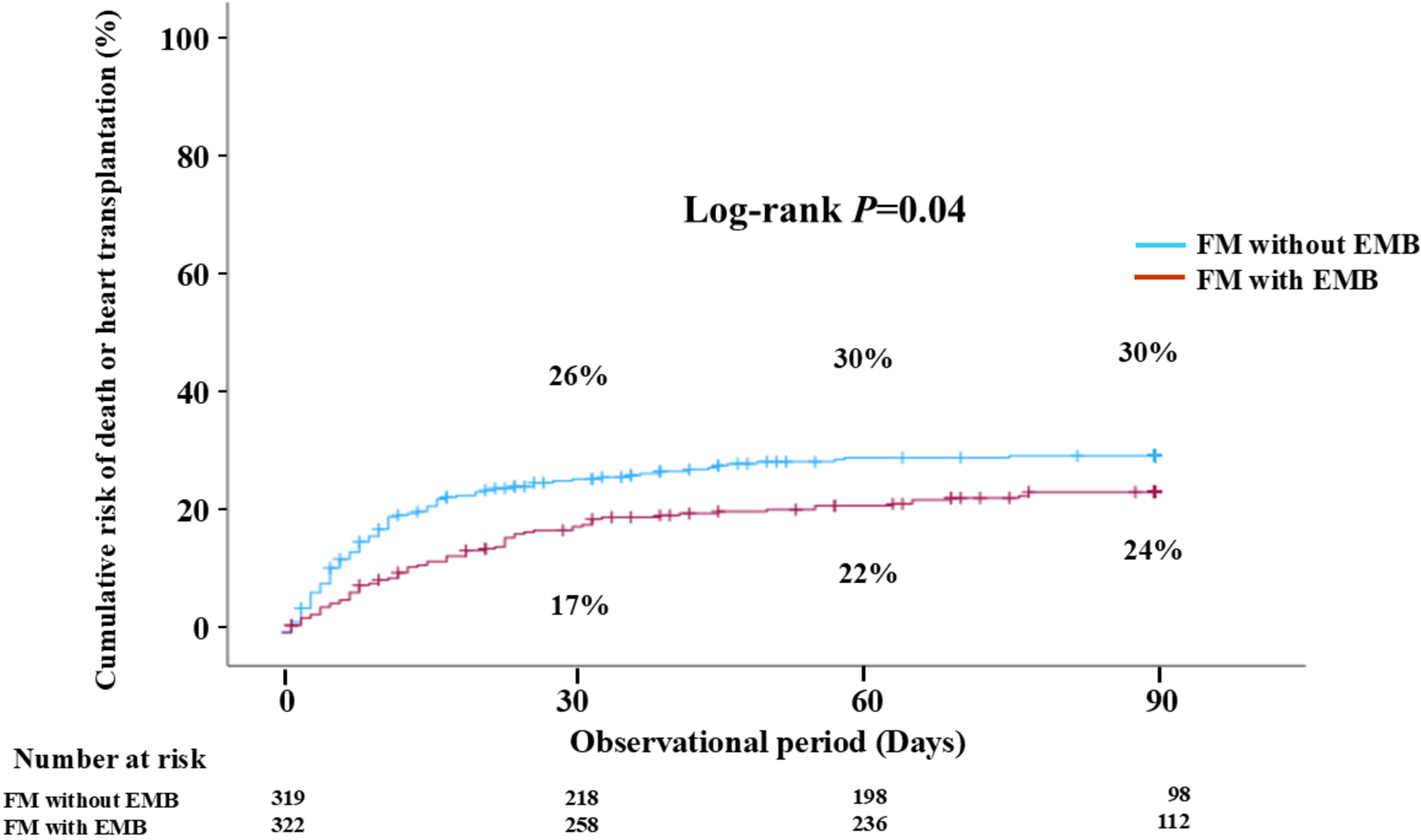
Ninety-day prognostic analysis according to endomyocardial biopsy in patients with fulminant myocarditis after consideration of immortal time bias. FM, fulminant myocarditis; EMB, endomyocardial biopsy.

### Factor Analysis Using In-hospital Variables between FM Patients with EMB and Those without

In performing EMB, the clinical state of the patients with FM, the cardiologist’s experience level, and the medical center’s level of expertise in EMB and cardiac pathology have been considered to be important factors.^13^ In addition to these factors, demographic data, factors of clinical severity, and medical therapies or interventions of the 655 patients comparing the 2 groups are shown in Table 2. Compared to FM patients without EMB, FM patients with EMB were significantly younger, had a higher incidence of cardiogenic shock on admission, and demonstrated a lower left ventricular ejection fraction. Regarding medical therapies and interventions, FM patients with EMB had a higher rate of initiation of MCS within 2 days of admission, and of administration of corticosteroids and immunoglobulins within 2 days of admission than FM patients without EMB. In addition, FM patients with EMB had a higher rate of introduction of GDMT for heart failure such as β-blockers, angiotensin-converting enzyme inhibitors/angiotensin II receptor blockers, and mineralocorticoid receptor antagonists, and higher total medical cost than those without EMB.

**Table 2.**
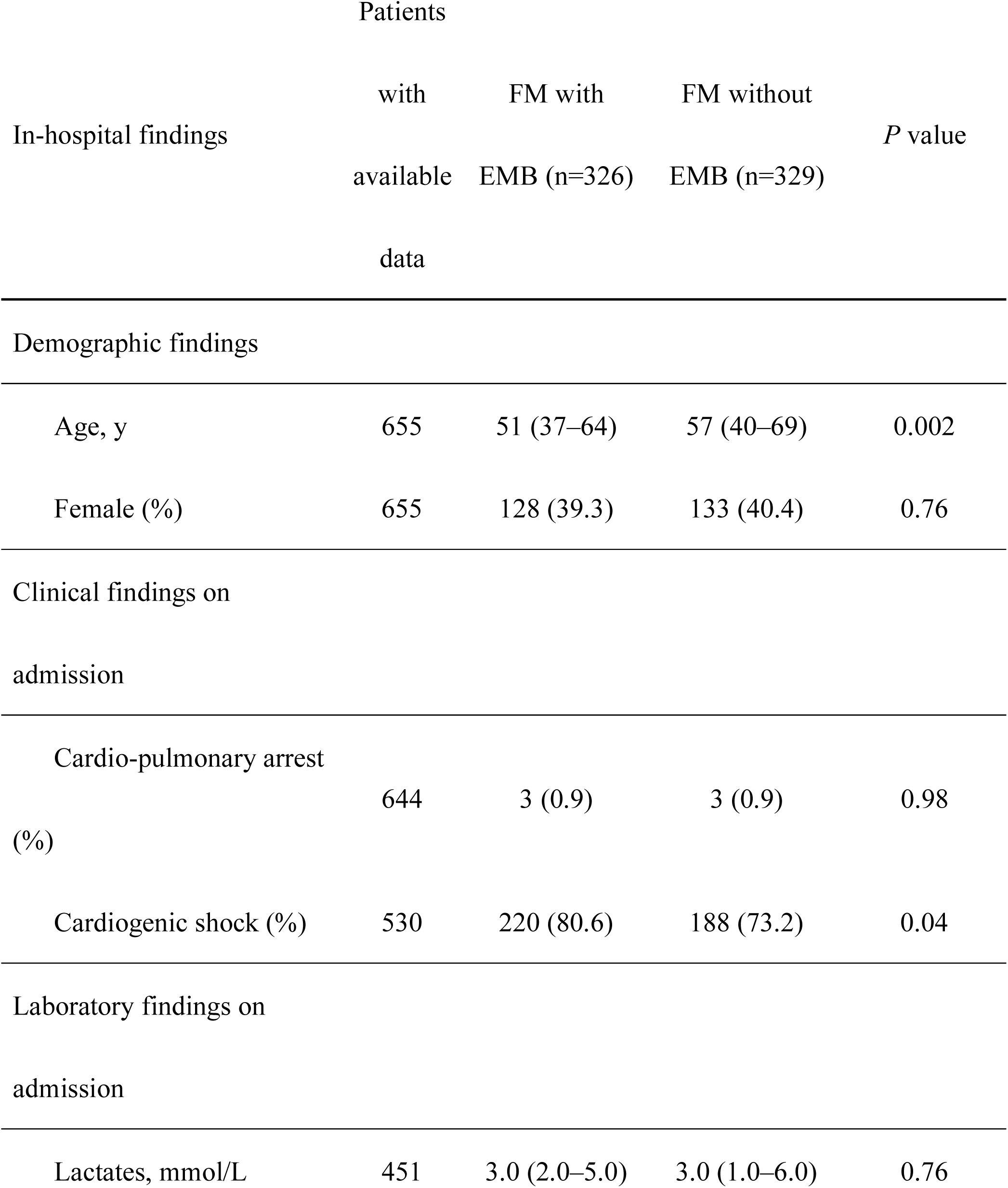
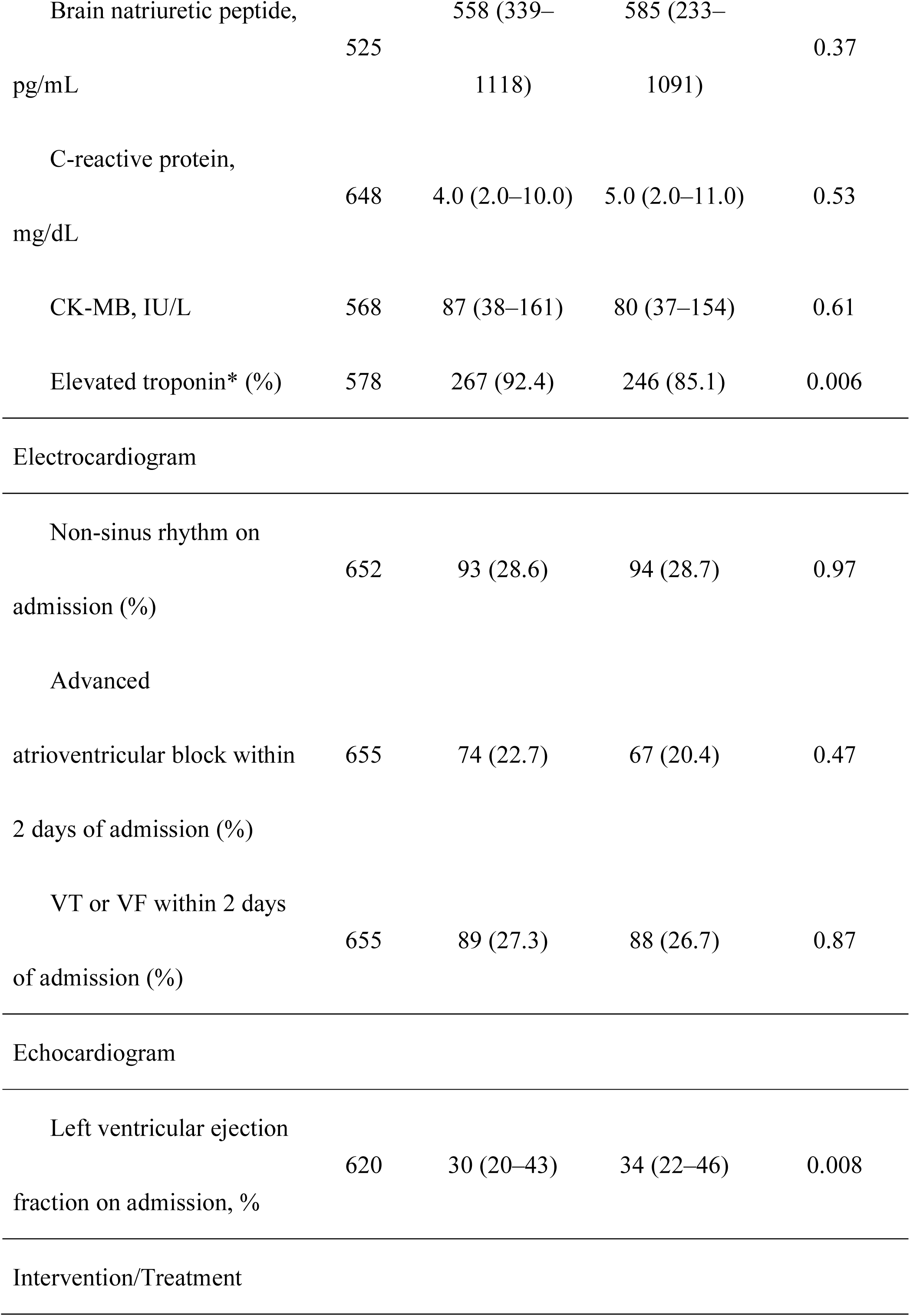
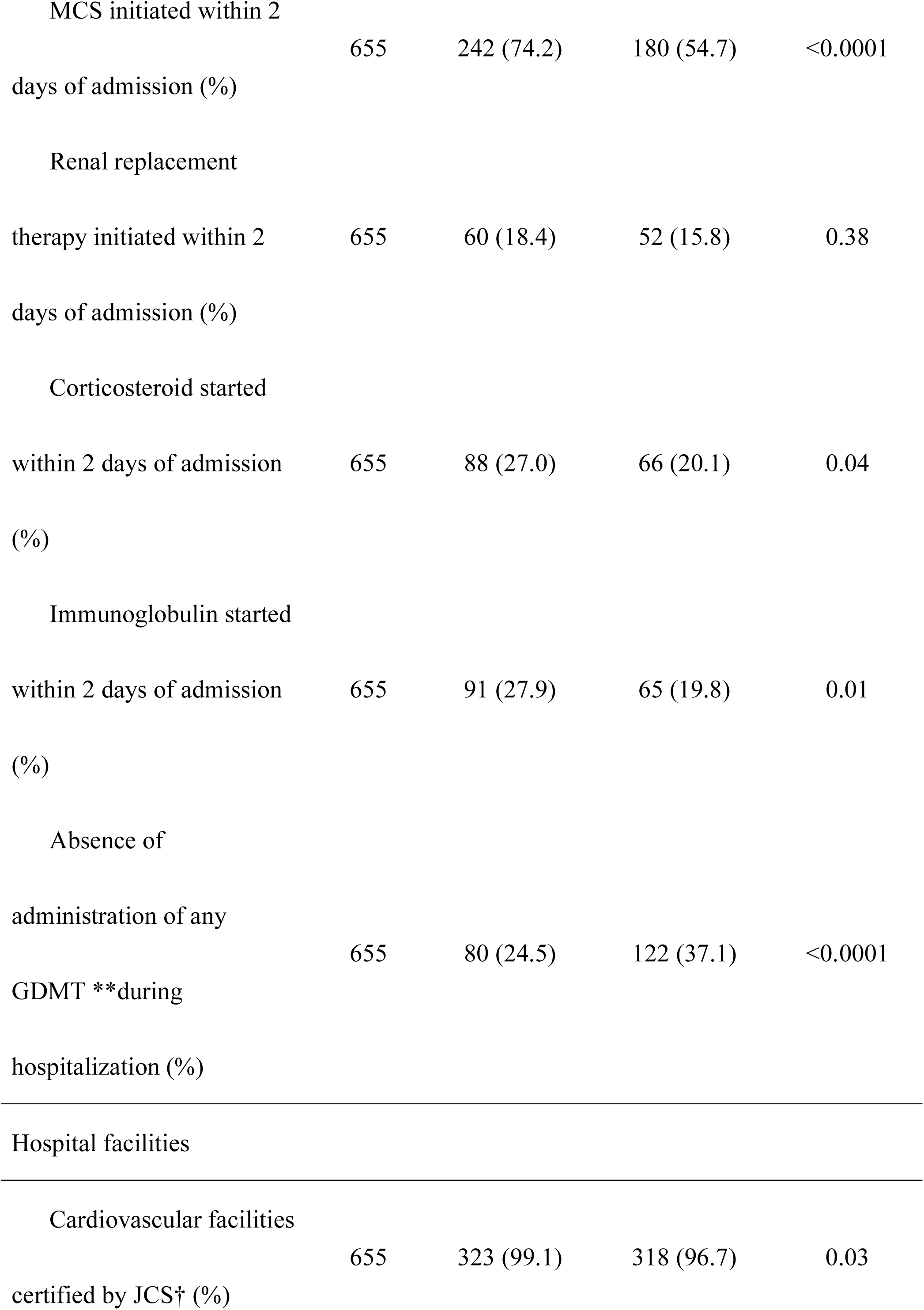
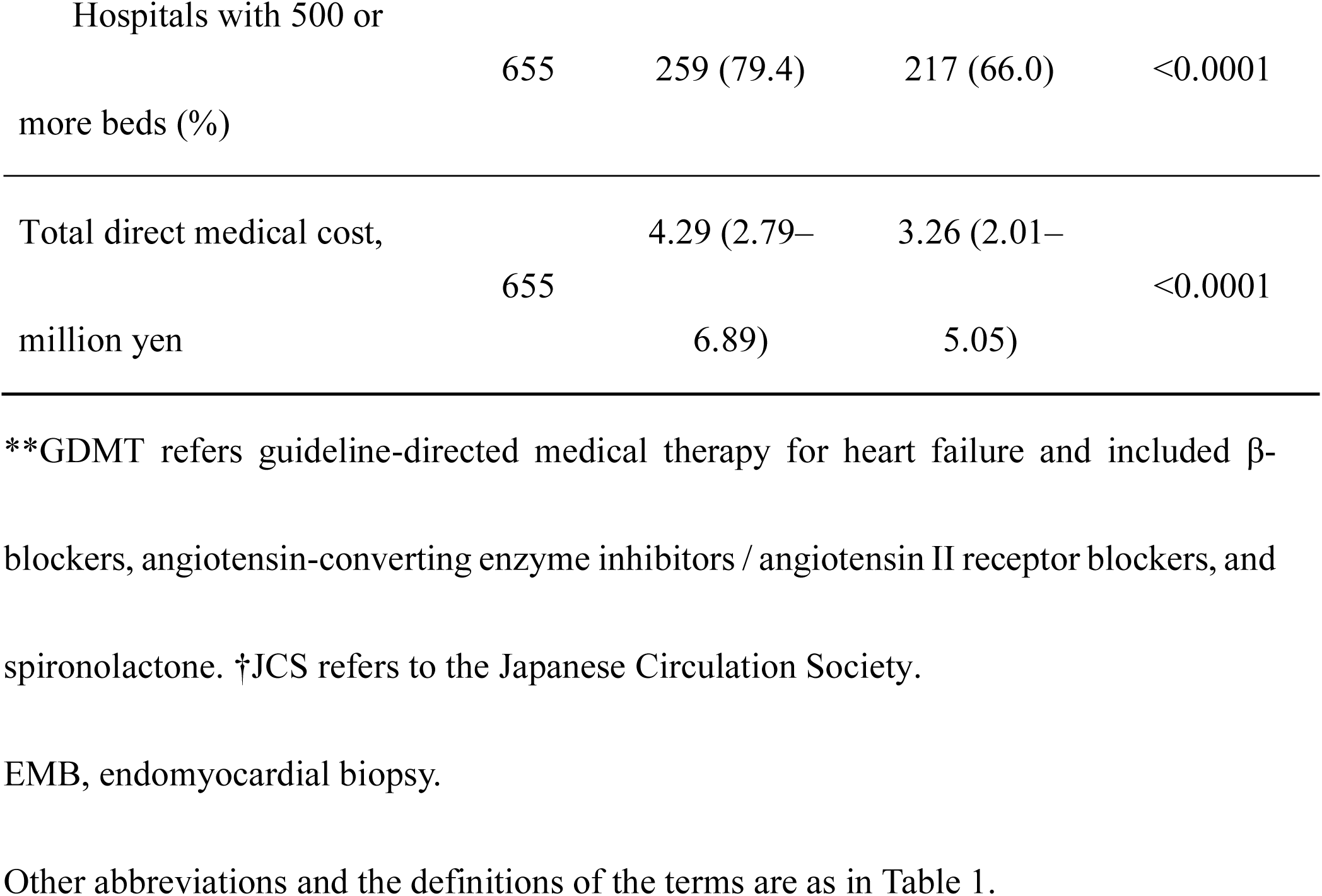
Comparison of Clinical Data between FM Patients with EMB and FM Patients without EMB.

Next, we performed a multivariable analysis of 90-day mortality. In all 655 patients, the absence of the administration of GDMT for heart failure was more strongly associated with a less favorable prognosis than age, non-sinus rhythm on admission, ventricular arrythmia within 2 days and left ventricular ejection fraction on admission of 40% or less; however, early initiation of MCS and early administration of immunomodulators were not significantly associated with 90-day mortality (Table 3).

**Table 3.** Ninety-Day Mortality Risk of Patients with FM (n=655)

| Variables | Univariable |  | Multivariable |  |
| --- | --- | --- | --- | --- |
|  | HR (95% CI) | *P* value | HR (95% CI) | *P* value |
| Age per 10 years | 1.34 (1.22–1.47) | <0.0001 | 1.34 (1.20–1.47) | <0.0001 |
| Female | 1.16 (0.85–1.56) | 0.35 | 1.18 (0.85–1.62) | 0.32 |
| Cardiogenic shock on admission | 0.71 (0.51–1.01) | 0.06 |  |  |
| Non-sinus rhythm on admission | 2.46 (1.82–3.33) | <0.0001 | 1.87 (1.34–2.62) | <0.0001 |
| Advanced AV block within 2 days of admission | 1.12 (0.79–1.60) | 0.53 |  |  |
| VT or VF within 2 days of admission | 2.15 (1.59–2.92) | <0.0001 | 1.42 (1.01–1.99) | 0.04 |
| LVEF < 40% on admission | 1.94 (1.31–2.86) | <0.0001 | 1.62 (1.08–2.42) | 0.02 |
| Corticosteroid started within 2 days of admission | 1.21 (0.86–1.69) | 0.22 |  |  |
| Immunoglobulin started<br>within 2 days of admission | 0.95 (0.66–1.35) | 0.76 |  |  |
| Absence of administration<br>of any GDMT* during<br>hospitalization | 4.59 (3.37–6.23) | <0.0001 | 4.34 (3.14–6.01) | <0.0001 |
| MCS initiated within 2<br>days of admission | 1.36 (0.98–1.89) | 0.07 |  |  |
| Cardiovascular facilities<br>certified by JCS† | 2.00 (0.50–8.06) | 0.33 |  |  |
| Hospital over 500 beds | 0.97 (0.69–1.35) | 0.84 |  |  |
| Total direct medical cost | 1.02 (1.00–1.04) | 0.02 | 1.01 (0.99–1.04) | 0.24 |

\*GDMT refers guideline-directed medical therapy for heart failure and included β-blockers, angiotensin-converting enzyme inhibitors / angiotensin II receptor blockers, and spironolactone. †JCS refers to the Japanese Circulation Society.
Other abbreviations and the definitions of the terms are as in Table 1.

## DISCUSSION

We performed a nationwide, large-scale, multicenter cohort study of patients with FM, involving 235 cardiovascular hospitals in Japan. We analyzed 736 clinical FM patients including 407 with EMB and 329 without EMB. The following key findings were observed: 1) FM patients who did not undergo EMB had a significantly higher mortality rate than FM patients who underwent EMB, after considering immortal time bias; 2) despite the high disease severity at the time of admission, FM patients who underwent EMB had a higher rate of initiation of MCS within 2 days of admission, a higher rate of administration of corticosteroids and immunoglobulins within 2 days of admission and a higher rate of introduction of GDMT for heart failure during hospitalization than FM patients who did not undergo EMB; and 3) the absence of GDMT for heart failure during hospitalization was more prevalent in FM patients who did not undergo EMB than those who underwent EMB and it was strongly associated with less a favorable prognosis.

Histological diagnosis is recommended for patients with FM because EMB is the only method that enables the accurate diagnosis and pathogenic clarification of myocarditis.^14^ EMB is highly crucial in determining the histological severity of inflammation and myocardial necrosis,^4^ and it can also provide information regarding virologic status of the myocardium when performed together with techniques such as immunohistochemistry, polymerase chain reaction analysis, and next generation sequencing.^15^ However, FM is a severe myocarditis characterized by hemodynamic instability requiring inotropes or MCS, and hence in real-world clinical practice, EMB is performed in a limited number of patients due to the perceived risks of the procedure, as well as physician inexperience or lack of appropriate systems in medical facilities.^13^ It has been reported that patients who underwent histological diagnosis by EMB had a more favorable prognosis than those who did not.^5,6^ However, the Kaplan-Meier curves in these studies showed a difference between the 2 groups from a very early phase. Considering that the analyses were performed at facilities where EMB could be performed, it is possible that the absence of EMB was associated with the high severity of the FM patients, and therefore, immortal time bias may have been present. The database in this study demonstrated that approximately 90% of all EMB were performed within 5 days of hospital admission and approximately 30% of all deaths occurred during the same period. For these reasons, we set the landmark of the survival curve as day 5 of admission, and performed prognosis analysis by classifying patients with and without EMB within 5 days of admission. Under the above conditions, FM patients without EMB had a significantly higher mortality rate within 90 days than FM patients with EMB.

On the other hand, there have been few reports to date that clearly identify the reasons or factors underlying the association between histological diagnosis by EMB and FM prognosis. Early histological diagnosis by EMB and early administration of immuno-modulation therapies may lead to an improved prognosis of FM.^5^ In the present study, approximately 90% of EMBs were performed within 5 days of admission; however, the early intravenous administration of corticosteroids or immunoglobulins started within 2 days of admission was not confirmed to be associated with an improvement of the prognosis of FM patients. Regarding the effects of immuno-modulation therapies, similar results were observed in another study using the same patient database.^16^ There is no strong evidence to support the use of immunosuppressive medicines including corticosteroids, and the level of this evidence has been low until now. The MYTHS trial will test the effectiveness of use the immunosuppressive medicines in FM patients in the United States (NCT05150704). The results of this trial will clarify the importance of histological diagnosis by EMB during the acute phase in FM patients.

There is no strong evidence to support the effectiveness of GDMT for heart failure in FM patients. β-blockers may be associated with a favorable prognosis in acute myocarditis patients owing to the prevention for ventricular arrythmia and sudden cardiac death.^17^ It has been reported that mineralocorticoid receptor antagonists demonstrated cardioprotective effects for viral myocarditis in an animal study.^18,19^ Angiotensin– converting coenzyme inhibitors and angiotensin II receptor blockers may be effective for myocardial injury associated with the treatment of severe acute respiratory syndrome coronavirus 2.^20^ In this study, the absence of administration of GDMT for heart failure was a strong prognostic factor on multivariate analysis. These results suggest that the important factor in determining the prognosis of the patients with FM is not the histological diagnosis by EMB itself, but rather the administration of at least one type of GDMT for heart failure. In other words, our results also suggest that histological diagnosis by EMB is valuable for guiding appropriate cardioprotective treatment for the patients with FM.

### Study Limitations

This study has several limitations. This retrospective study was conducted on Japanese patients in institutions in Japan, and clinical practices may differ from those of other countries. However, the study is similar to other studies regarding baseline characteristics, treatment patterns including indication for MCS, and mortality rate.^9^ Second, the FM patients who experienced cardiogenic shock but did not undergo catecholamine or temporary MCS were not included in this study. Third, the administration of GDMT for heart failure might have been influenced by the severity of the FM patient’s condition, such as severe left ventricular dysfunction or impaired blood flow to multiple organs. In addition, there is a lack of data on the administration of angiotensin receptor-neprilysin inhibitor and sodium-glucose cotransporter 2 inhibitors.^21^ These two medicines were not covered by health insurance in Japan at that time. Future studies are needed to determine the efficacy of these two medicines in patients with FM. Fourth, this study did not include sufficient information on virus genome analysis of EMBs because it is not routine clinical practice in Japan. It is still controversial whether viral genome sequencing is beneficial for the diagnosis and treatment of myocarditis.^22^ Parvovirus 19 is frequently detected in virus-positive lymphocytic FM patients and immunosuppression does not exacerbate parvovirus 19 replication in patients with myocarditis.^23,24^ Virus genome analysis might have enabled patients with myocarditis to undergo specific treatments.

## Conclusions

In conclusion, we demonstrated the influence of histological diagnosis by EMB in the early phase after admission on advanced medical treatment and prognosis in patients with FM. Database analysis with careful consideration of immortal time bias demonstrated that performing the histological diagnosis by EMB in the early phase after admission had a prognostic benefit. GDMT for heart failure may be more effective than early initiation of immuno-modulation therapies or early initiation of MCS in improving the prognosis of patients with FM. Comprehensive patient management, including accurate histological diagnosis by EMB and appropriate cardioprotective therapy for heart failure may determine the prognosis of patients with FM.

## Data Availability

All of the data mentioned in the manuscript will not be shared.

## Acknowledgments

The authors gratefully acknowledge the medical editors of the Center for International Education and Research of Tokyo Medical University for English editing of the manuscript, and also acknowledge Dr. Sho Komukai, Department of Data Health Science of Tokyo Medical University for providing technical advice regarding the statistical analysis.

## Institutional review board information

This study was approved by the Institutional Review Boards in Tokyo Medical University (study registration number: T2020–0041), Nara Medical University (study registration number: 2256), and the Japanese Circulation Society (study registration number: 10).

## Nonstandard Abbreviations and Acronyms

EMB: endomyocardial biopsy
FM: fulminant myocarditis
GDMT: guideline-directed medical therapy
HTx: heart transplantation
JCS: the Japanese Circulation Society
JROAD–DPC: the Japanese Registry of All Cardiac and Vascular Diseases–Diagnosis Procedure Combination
MCS: mechanical circulatory support

